# Immunity status in patients with periprosthetic knee joint infection

**DOI:** 10.1101/2021.08.23.21261316

**Authors:** Irina A. Mamonova, Irina. V. Babushkina, Aleksandr S. Bondarenko, Vladimir Yu. Ulyanov

## Abstract

Implant-associated infection is one of the most severe complications after total replacement of larger joints. The diagnosis of periprosthetic infection are often impeded as 5-34% of bacteriological tests are false-negative. Now the investigations of the immune status of patients suspected of periprosthetic infection are considered perspective as these are aimed at finding the most significant indicators for underlying the diagnosis of this pathology. In this research, we assessed immunology indicators in peripheral blood of patients with periprosthetic infection in their knee joints. We determined immunology characteristics associated with the clinical aspects of the progressing pathology. Our findings can be used as the basis for further search of pathophysiological mechanisms of periprosthetic infection progress in the knee joint as well as defining new diagnostic approaches.

**Reviewers:**

Ivanov A.N., MD, DSc;

Prof. Puchinyan D.M., MD, DSc

## Background

**I**mplant-associated infection ranges with the most severe complications of larger joint replacements. Periprosthetic infection is often difficult to diagnose, its pathogenesis is underlain by the microorganism ability to form microbe biofilms on the surface of the implanted joints hindering the isolation of the agent from specimens for examination [1]. 5 to 34 percent of patients with periprosthetic infection receive false-negative results to their bacteriological tests. Nowadays the examination of the immune status in patients suspected of having periprosthetic infection is getting promising as it’s aimed at determining the most relevant indicators that may form the basis for the diagnostics of this pathology [2, 3].

Therefore the research was aimed at the investigation of the immune status in patients with periprosthetic infection in their knee joints to define the most relevant diagnostic indicators.

## Material and methods

The study involved 20 patients under treatment in Scientific Research Institute of Traumatology, Orthopedics and Neurosurgery, Federal State Budgetary Educational Institution of Higher Education ‘V.I. Razumovsky Saratov State Medical University’ (15 women and 5 men) with suppurative complications after surgical interventions. The mean age of the patients was 63.4 (55.6; 69.8). The study excluding criteria were systemic disease such as rheumatoid arthritis, ankylosing spondylitis, it also didn’t involve patients with severer comorbidity – diabetes mellitus, cardiac or renal insufficiency and other.

The control group was made up of 20 apparently healthy volunteers (12 women and 8 men) aged 63.4 (55.3; 68.1). The examined individuals had no musculoskeletal pathologies, allergic, autoimmune, infectious inflammatory diseases and were hepatitis B, C or HIV negative.

Lymphocyte typing in peripheral blood was run with laser flow cytometry method in BD FACS Canto II (BD, USA). The lymphocytes and the pattern of their subset populations were identified with BD Multitest 6-Color TBNK Reagent (BD, USA) direct immunofluorescence kit. T-lymphocytes (CD3+), T-helpers (CD3+CD4+), cytotoxic T-lymphocytes (CD3+CD8+), B-lymphocytes (CD3-CD19+), natural killers (CD3-CD16+CD56+), natural killers/T-lymphocytes (CD3+CD16+CD56+) were identified with phycoerythrin (PE), fluorescein isothiocyanate (FITC), peridinin-chlorophyll-protein (Per-CP) or allophycocyanin (APC) labeled monoclonal antibodies. The absolute and relative indicators of lymphocyte populations were calculated in BD Trucount (BD, USA) test tubes.

The humoral component of immune system was determined with the number of serum A, M, G immunoglobulins by immunoturbidimetric method in Sapphire-400 automated open biochemical analyzer (Hirose Electronic System Co., Japan) with Immunoglobulin A FS, G FS, M FS panels (DiaSys Diagnostic Systems GmbH, Germany) according to instructions provided with the kit.

Serum cytokines (TNFα, IL-4, IL-6, IL-10) levels were found with enzyme-linked immunoelectrodiffusion essay with diagnostic systems Interlejkin-4-IFA-BEST, Interlejkin-6-IFA-BEST, Alfa-FNO-IFA-BEST(Vector-Best Ltd., Russia) in Epoch (Biotek, USA) device according to the instruction in the kit.

The findings were processed in Microsoft Excel 2016 and Statistica 13 Trial software suites. The Shapiro-Wilk test was used to check the normality of quantitative indicators distribution. The statistical analysis involved nonparametric method and the calculation of mean (M), standard deviation (SD), median (Me), 25^th^ and 75^th^ quartiles (Q). The statistical significance was evaluated with Mann-Whitney U-test.

The principal component analysis was run using SPSS (11-2018) software package. The applicability of data for factor analysis and the significance of the derived factor model were verified with Kaiser-Meyer-Olkin measure of sampling adequacy (the values should be within 0.5 to 1 range) and the Bartlett’s test (the model is analyzable if p<0.05). The factoring was run using principal component analysis and the best factor composition was found though Varimax rotation method with Kaiser normalization.

## Results

We examined the immune systems of patients with periprosthetic infection developed after surgical intervention for total knee replacement.

The cell immunity findings is patients with periprosthetic infection was compared to the controls (Table 1). The statistically significant decrease (p=0.001) in the absolute number of T-lymphocytes (CD3+CD19-) by T-helpers (p=0.001) as well as T-suppressors (p=0.015) was observed in peripheral blood of patients with periprosthetic infection. However no statistically significant change in CD4/CD8 lymphocyte subset populations was detected. Helper-suppressor cell ratio in the controls was 1.5 (1.3; 1.7) while in the patients with periprosthetic infection it was 1.4 (1.3; 1.7).

**Table 1.** Immune status indicators in patients with periprosthetic infection of knee joints.

| Indicators | Control patients (n=20) | Patients with periprosthetic infection of knee joints (n=20) |
| --- | --- | --- |
| CD3+CD19-, abs | 1781.0 (1433.8;1808.9) | 1177.8 (1091.0; 1262.6) * |
| CD3+CD19-, % | 76.6 (71.3; 78.9) | 72.4 (69,3; 79,7) |
| CD3+CD8+, abs | 628.7 (549.9; 727.3) | 491.3 (250.3; 589.7) * |
| CD3+CD8+, % | 27.3 (24.0; 32.2) | 25.1 (18.1; 29.8) |
| CD3+CD4+, abs | 997.9 (959.3; 1123.8) | 653.0 (606.2; 801.0) * |
| CD3+CD4+, % | 43.7 (41.8; 46.9) | 40.7 (37.6; 46.6) |
| CD3-CD16+CD56+, abs | 302.1 (123.5; 356.9) | 174.6 (155.5; 251.8) * |
| CD3-CD16+CD56+, % | 13.3 (6.4; 17.8) | 10.5 (9.4; 15.4) |
| CD3-CD19+, abs | 307.1 (305.0; 17.8) | 177.6 (120.8; 280.5) * |
| CD3-CD19+, % | 13.3 (13,4; 14.5) | 12.9 (8.0; 17.9) |
| IgA (pg/ml) | 106.0 (95.0; 138.5) | 177.5 (132.0; 208.0) * |
| IgM (pg/ml) | 153.6 (128.0; 182.5) | 108.0 (86.0; 136.0) * |
| IgG (pg/ml) | 1652.2 (1564.5; 1766.0) | 1362.0 (1161.0; 1613.0) * |
| TNFα (pg/ml) | 4.9 (4.5;6.7) | 19.3 (14.0; 23.7)* |
| IL-4 (pg/ml) | 0.2 (0.1; 0.3) | 0.2 (0.1; 0.2) |
| IL-6 (pg/ml) | 7.1 (5.8; 7.7) | 26.8 (23.4; 36.4)* |
| IL-10 (pg/ml) | 16.8 (12.6;21.7) | 9.1 (6.3; 9.3) * |

Note: \* – statistically significant differences as compared to the controls

We established a statistically significant decrease (p=0.032) in the absolute number of NK-cells (CD3-CD16+CD56+) as compared to the group of apparently healthy donors.

We studied indicators of humoral immune response in peripheral blood of patients with periprosthetic knee infection progressing after total replacement (Table 1). The analysis of the findings revealed statistically significant decrease in the absolute number of B-lymphocytes (CD3-CD19+) in peripheral blood of patients with periprosthetic infection (p=0.029) as compared to the group of apparently healthy volunteers (Table 1). The level of serum immunoglobulin A presented the rise (p=0.001) and the concentrations of immunoglobulin M (p=0.001) and G (p=0.001) classes decreased.

Then we compared cytokine profiles of apparently healthy individuals and patients with periprosthetic infection (Table 1), and found the increased level of proinflammatory cytokines TNFα (p=0.001) and IL-6 (p=0.001) while the concentration of anti-inflammatory cytokine IL-10 (p=0.001) in blood serum was decreased suggesting the compromised state of immune system in patients with periprosthetic infection as well as active inflammation process around the replaced joint. No statistically significant difference in serum IL-4 content between the patients with periprosthetic infection and relatively healthy volunteers was found.

The factor analysis of the immunogram indicators in patients with periprosthetic infection of their knees (Table 2) enabled establishing the diagnostically relevant indicators reflecting the progressing pathology and designing the factor matrix. The homogeneity was achieved by the elimination of out-of-range data average ± 2 mean quadratic deviations. As the data resulted from valid tests and featured high reliability we considered possible to analyze the moderate-sized samples. The feasibility of data for factor analysis and the significance of the resulted factor model were proven with Kaiser-Meyer-Olkin measure as well as the Bartlett’s test. The analysis led to determining the two factors (KMO=0.513 and the Bartlett’s test=0.001<0.05) accounting for a total of 77.6 percent of the changes in indicators.

**Table 2.** The matrix of the rotated immune indicator factors in patents with periprosthetic knee infection.

| Indicators | Factor |  |
| --- | --- | --- |
|  | 1 | 2 |
| Immunoglobulin M (IgM) | 0.922 | - |
| Tumor necrosis factor (TNFα) | 0.917 | - |
| Interleukin-10 (IL-10) | -0.521 | - |
| Absolute number of NK-cells (CD3-CD16+CD56+) | - | -0.923 |
| Absolute number of cytotoxic T-lymphocytes (CD3+CD8+) | - | 0.916 |
| Characteristic values | 2.209 | 1.673 |
| % of explained variance | 44.171 | 33.462 |

The first factor comprises 44.2 percent of all possible states and includes indicators of humoral immunity and cytokine profile of the patients with implant-associated infection. We demonstrated strong positive correlation of serum IgM concentration (factor loading of the variable +0.922) with TNFα (+0.917) and average correlation (−0.512) with IL-10 concentration. The second factor made 33.4 percent of the total dispersion and comprised cell immunity indicators. The decrease in natural killer (CD3-CD16+CD56+) content correlated with the increase in cytotoxic T-lymphocytes (CD3+CD8+). The elements of the second factor correlated with the absolute indicators of cell immunity only.

## Discussion

**O**ur factor analysis detected immune characteristics associated with clinical aspects of the pathology. The implant associated infection features low content of immunoglobulin M suggesting the deficiency of primary humoral response to constantly increasing antigen load. However the decrease in serum IL-10 possibly suggests the deficit of B-cell stimulation facilitating the decrease in their proliferation and immunoglobulin output. The revealed negative correlation of TNFα and IL-10 indicators refers the developed imbalance of pro- and anti-inflammatory cytokines. The abnormality of cell immunity was thus established. The analysis also revealed the invert correlation of T lymphocytes (CD3+CD8+) and NK cells (CD3-CD16+CD56+) in peripheral blood. Killer lymphocytes are known to eliminate foreign or altered macroorganism cells in the absence of molecules of class I major histocompatibility complex (MHCI) on their surface despite the contents of antibodies or complement system. We observed the decrease in NK cells in isolates of patients with periprosthetic infection. Moreover, the decrease in T-helpers and T-suppressors subpopulations was found in peripheral blood though no significant changes in immunoregulatory index was detected suggesting the normal balance of cells in the population as well as the absence of dominance of some cells over others. The low content of T-helpers and T-suppressors subpopulations was related to the decrease of absolute T lymphocytes count in peripheral blood.

## Conclusion

**T**he results of the research can form the basis for further investigation on pathophysiology mechanisms of periprosthetic infection progress in knee joint as well as the search for new diagnostic approaches.

## Data Availability

The data that support the findings of this study are available from the corresponding author upon reasonable request

